# Epidemic Trend Analysis of SARS-CoV-2 in SAARC Countries Using Modified SIR (M-SIR) Predictive Model

**DOI:** 10.1101/2020.06.29.20142513

**Authors:** Samrat Kumar Dey, Md. Mahbubur Rahman, Kabid Hassan Shibly, Umme Raihan Siddiqi, Arpita Howlader

**Affiliations:** Department of Computer Science and Engineering (CSE), Dhaka International University (DIU), Dhaka-1205, Bangladesh; Department of Computer Science and Engineering (CSE), Military Institute of Science and Technology (MIST), Mirpur Cantonment, Dhaka-1216, Bangladesh; Department of Physiology, Shaheed Suhrawardy Medical College (ShSMC), Dhaka-1207, Bangladesh; Department of Computer and Communication Engineering (CCE), Patuakhali Science and Technology University (PSTU), Dumki-8602, Patuakhali, Bangladesh

**Keywords:** SARS-CoV-2, COVID-19, epidemics, SAARC, modified susceptible-infected-recovered, prediction model

## Abstract

A novel coronavirus causing the severe and fatal respiratory syndrome was identified in China, is now producing outbreaks in more than two hundred countries around the world, and became pandemic by the time. In this article, a modified version of the well known mathematical epidemic model Susceptible (S)- Infected (I)- Recovered (R) is used to analyze the epidemic’s course of COVID-19 in eight different countries of the South Asian Association for Regional Cooperation (SAARC). To achieve this goal, the parameters of the SIR model are identified by using publicly available data for the corresponding countries: Afghanistan, Bangladesh, Bhutan, India, the Maldives, Nepal, Pakistan and Sri Lanka. Based on the prediction model we estimated the epidemic trend of COVID-19 outbreak in SAARC countries for 20 days, 90 days, and 180 days respectively. An SML (short-mid-long) term prediction model has been designed to understand the early dynamics of COVID-19 Epidemic in the south-east Asian region. The maximum and minimum basic reproduction number (R_0_ = 1.33 and 1.07) for SAARC countries are predicted to be in Pakistan and Bhutan. We equate simulation results with real data in the SAARC countries on the COVID-19 outbreak, and model potential countermeasure implementation scenarios. Our results should provide policymakers with a method for evaluating the impacts of possible interventions, including lockdown and social distancing, as well as testing and contact tracking.

## 1. Introduction

A novel coronavirus (SARS-CoV-2), called COVID-19, caused an outbreak in the city of Wuhan, Hubei Province, China that is linked to the Huanan Seafood Wholesale Market ^**1, 2**^ in late December 2019. Till now, more than 200 countries around the world have been infected by a novel coronavirus. As of May 30, 2020, according to the World Health Organization (WHO), globally 5817385 confirmed cases have reported with a death count of 362705^**3**^. Among them, the south-east Asia region has confirmed 4.2% of cases while the case fatality ratio is 2.86%.

The very first COVID-19 patient was treated for being coronavirus positive in Wuhan, China, at the beginning of December 2019. In the south-east Asia region, the very first coronavirus infected patient was detected in Pakistan on February 26, 2020^**4**^. On 30 January 30, 2020, WHO officially declared this outbreak of COVID-19 as global pandemic^**5**^. To mitigate the spread of COVID-19, the affected countries of the world have taken various measures, including citywide lockdown, social distancing, traffic halt, community management, and information on health education knowledge. More importantly, the outbreak of COVID-19 possessed a massive threat to global health and economics all over the world. One of the significant feature of novel coronavirus unlike other infectious diseases like SARS (Severe Acute Respiratory Syndrome), and MERS (Middle East Respesoritory Syndrom), it causes asymptomatic infections (symptoms are very mild)^**6,7, 8**^. In this circumstances, the rate of transmission among a large number of people can increase within no time. According to the latest World Health Organization survey, only 87.9% of COVID-19 patients have a fever, and 67.7% have dry cough^**9**^. Therefore, this is highly crucial to estimate the intensity of the COVID-19 epidemic and predict the time course, peak time, total duration, and so on. Our study focuses on the COVID-19 case prediction using a modified SIR model (M-SIR) in the countries of the South Asian Association for Regional Cooperation (SAARC). SAARC is considered as an intergovernmental organization and geopolitical union of states in South Asia. Its member states are Afghanistan, Bangladesh, Bhutan, India, the Maldives, Nepal, Pakistan and Sri Lanka. Our aim is to develop a prediction model for the SAARC countries to understand the epidemiological trend of novel coronavirus outbreak in these countries. Here we explore a modified version based on the Susceptible (S)- Infected (I)- Recovered (R) epidemic model to predict the short term (20 days), mid-term (90 days) and long term (180 days) evaluation of COVID-19 situation in these countries of SAARC regions.

## 2. Methodology

### 2.1 Dataset Analysis

For this paper, we have used datasets from various sources for our analysis and building the model. We have used four different sources of the dataset including, COVID-19 Data Repository by the Center for Systems Science and Engineering (CSSE) at Johns Hopkins University (January - May 2020)^**10**^, COVID-19 Dataset (January - May 2020)^**11**^, COVID-19 Open Research Dataset (CORD-19)^**12**^ and population by country 2020 dataset^**13**^. **Table 1** provides insight on each dataset and their respective data files with their column description.

**Table 1:** Tabular representation of different data sources of COVID-19

| Dataset | Description | Columns |
| --- | --- | --- |
| **Novel Coronavirus 2019 Dataset** |  |  |
| COVID19_line_list_data.csv | This file is an aggregated version of the Novel Coronavirus dataset collected by Johns Hopkins University. | Id, case_in_country, reporting date, summary, location, country, gender, age, symptom_onset, If_onset_approximate, hosp_visit_date, exposure_start, exposure_end, visiting Wuhan, from Wuhan, death, recovered, symptom, source |
| covid_19_data.csv | Daily level information on the number of COVID-19 affected cases across the globe | Sno, Observation Date, Province/State, Country/Region, Last Update, Confirmed, Deaths, Recovered |
| time_series_covid_19_confirmed.csv | Time series data on the number of confirmed cases | Province/State, Country/Region, Lat, Long, 1/22/20...5/6/20 |
| time_series_covid_19_deaths.csv | Time series data on the number of death cases | Province/State, Country/Region, Lat, Long, 1/22/20...5/6/20 |
| time_series_covid_19_recovered.csv | Time series data on the number of recovered cases | Province/State, Country/Region, Lat, Long, 1/22/20...5/6/20 |
| **COVID-19 (nCoV-19) Coronavirus Spread Dataset** |  |  |
| covid_19_clean_complete.csv | The file contains the cumulative count of confirmed, death and recovered cases of COVID-19 from different countries from January 2020 | Province/State, Country/Region, Lat, Long, Date, Confirmed, Deaths, Recovered |
| **COVID-19 Open Research Dataset** |  |  |
| metadata.csv | This dataset contains metadata for 59k articles on COVID-19 | Cord_uid, sha, source_x, title, doi, pmcid, license, abstract, publish_time, authors, journal |
| **Population by country 2020 dataset** |  |  |
| population_by_country_2020.csv | This dataset contains the information from 235 countries along with their population till 2020. | Country (or dependency), population (2020), Yearly Change, Net Change, Density (P/Km<sup>2</sup>), Land Area (Km<sup>2</sup>), Migrants (net), Fert. Rate, Med. Age, Urban Pop %, World Share |

We also developed a custom dataset to develop and evaluate our model. The following **Table 2** is developed based on collected data from different sources.

**Table 2:**
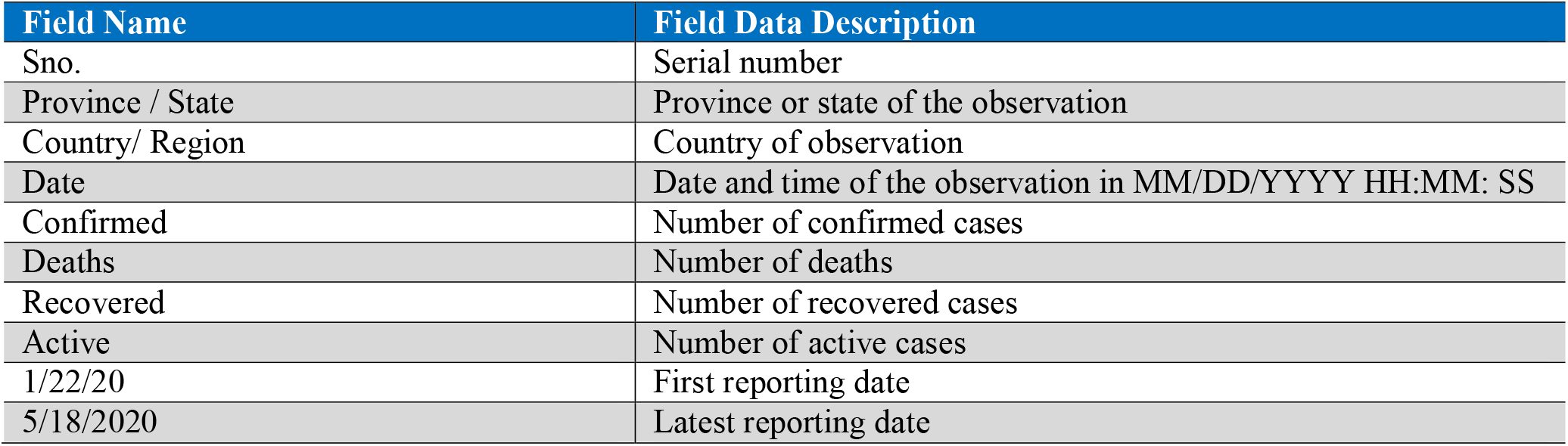
Columns description of custom build COVID-19 dataset

### 2.2 Modified SIR (M-SIR) Model

Predictive mathematical disease models are important for understanding the trajectory of the outbreak and for preparing successful response strategies. One commonly used model is the human-to-human transmission SIR model, which defines people’s flow through three mutually exclusive stages of infection: Susceptible (S), Infected (I), and Recovered (R). Most epidemic models are based on dividing the population into a small number of sections. Each person is identical in terms of their status with the considered disease. The SIR model based on three sections: Susceptible (S) is the class for those who are susceptible to infection. This can include passive immune systems as soon as they lose their immunity. In the Infected (I) class, the parasite level within the host is large enough, and there is a possibility of spreading the infection to other susceptible people. The Recovered (R) class includes all infected and recovered individuals. This epidemiological model captures the dynamics of acute infections that, after recovery, confer lifelong immunity. In general, the overall size of the population is considered constant N=S+I+R. The two cases should be examined and characterized by the inclusion or exclusion of demographic factors. Let’s assume in the SIR model; there is a natural host lifespan of 1 / µ years. Then the rate at which individuals in an epidemiological compartment suffer from natural mortality is given by µ. It is important to emphasize that this factor is independent of the disease and should not reflect the pathogenicity of the infectious agent. Diachronically, it can be said,

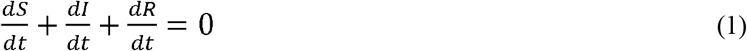

So, the SIR model can be defined as,

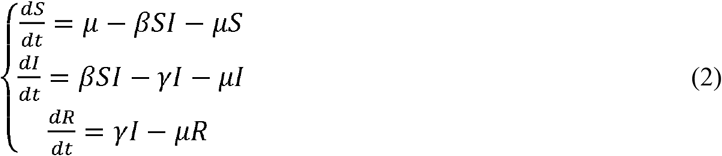

Here, the initial conditions **S (0) > 0, I (0)** ≥ **0 and R (0)** ≥ **0**. It is important to enter the expression of basic reproduction number ***R***_**0**_ for this model. The basic reproduction number ***R***_**0**_ is the parameter that estimates whether a disease has spread to the population or not. If the estimated ***R***_**0**_ < 1, we can assume that the disease will die out, and if ***R***_**0**_ = 1, the disease remains in the system and is stable. But if ***R***_**0**_ > 1, the disease will spread and cause an outbreak. The higher the value of ***R***_**0**_, the more difficult it is to control.

### 2.3 Proposed Model and Algorithm

In this exploration, we have used a susceptible (S)-infected (I)-recovered (R) epidemic model. In a general SIR model, transmission rate (β) and recovery rate (γ) are considered as two time-invariant variables. Moreover, several research studies have shown that a SIR model works much better in presenting the information contained in the confirmed case data than an SEIR model^**14**^. Therefore we have developed a model that can dynamically adjust the crucial parameters while working on time-varying data, which is also treated as a modified SIR (M-SIR) model ^**15**^. However, in a basic SIR model, the reproduction number (R) is a simple division of transmission and recovery rates, as shown in Equation (3).

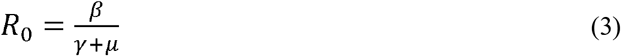

For building the model, we modified the primary reproduction number, R0with respect to time (t). Equation (4) represents the changes that happened depending on time (t).

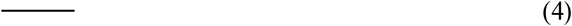

We also considered detectable () and non-detectable (infected persons for building our model effectively, as shown in Equation (5).

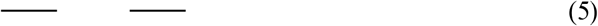

In general, detectable persons contain a lower transmission rate than non-detectable persons. Therefore we calculated the transmission rate (β) and recovery rate (γ) for each country of the SAARC region. For instance, Bangladesh has 165 million people^**16**^, and the first confirmed case reported in the country on March 7, 2020. As of May 30, 2020, a total of 42,844 COVID-19 infected people detected in the country^3^. Depending on the modified SIR model, Bangladesh contains a transmission rate of 0.63 with a recovery rate rate of 0.49. The initial reproduction number for the country is 1.27. Based on available data sources of every SAARC country, we have calculated these parameters for the prediction model, which is shown in **Table 3. Algorithm 1** represents the working procedure of our proposed (M-SIR) prediction model.

**Table 3.** Transmission rate, Recovery rate, and Reproduction number of SAARC countries.

| Countries | Transmission rate ($\beta$) | Recovery rate ($\gamma$) | Reproduction Number ($R_0$) |
| --- | --- | --- | --- |
| Afghanistan | 0.5997691 | 0.4889147 | 1.22673566575 |
| Bangladesh | 0.6286615 | 0.4932285 | 1.2745847006 |
| Bhutan | 0.5334682 | 0.4982385 | 1.07070850607 |
| India | 0.5060977 | 0.3955793 | 1.27938367857 |
| Maldives | 0.5774223 | 0.4920300 | 1.17355100299 |
| Nepal | 0.5406592 | 0.4954823 | 1.09117762632 |
| Pakistan | 0.5254487 | 0.3947732 | 1.33101411139 |
| Sri Lanka | 0.5595021 | 0.4933888 | 1.13399838018 |

#### Algorithm 1: M-SIR prediction model

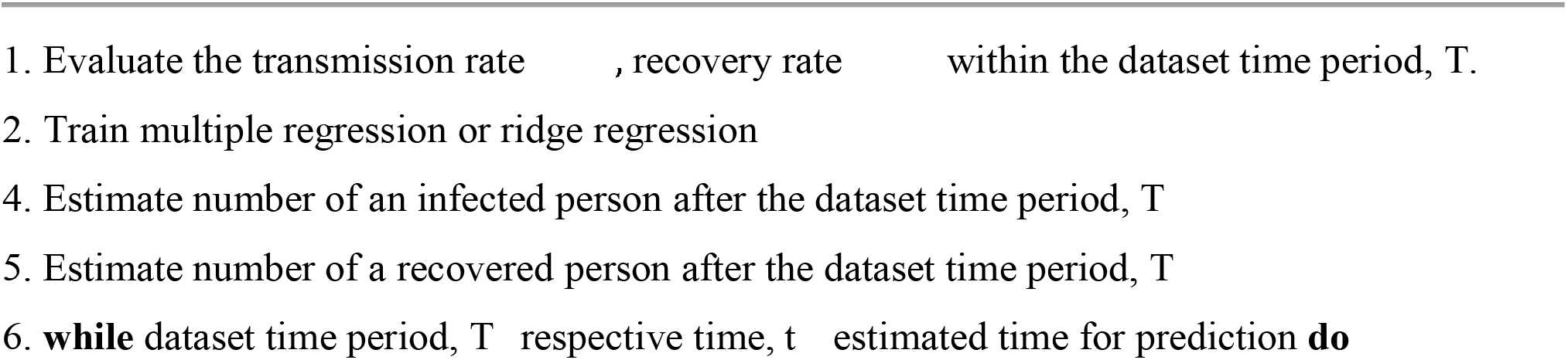
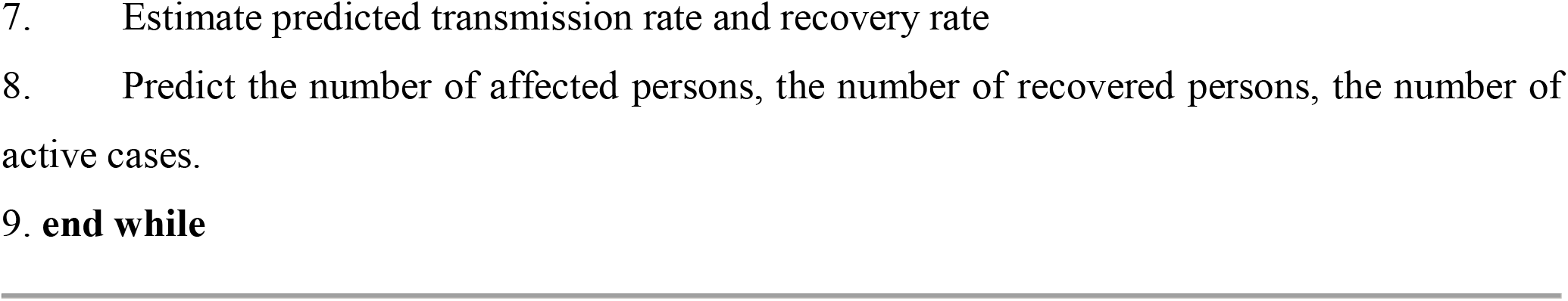

## 3. Results

We performed our simulations and tabulated the predicted results (short term, midterm, and long term) with all countries from the SAARC region. We also explored 3D parameter space using gradient descent to minimize the error. However, a lag parameter is also used in this experiment to reduce the gap of first confirmed cases of SAARC countries’ pandemic situation. The following **Table 3** represents the transmission rate (β), recovery rate (γ), and the reproduction numbers (*R*_0_) of all countries in the SAARC regions. Bangladesh contains the highest transmission rate of (β = ∼0.63) in the SAARC region, whereas India shows a much lower transmission rate of only ∼0.51 comparing with other countries. Also, data analysis visualization tool have been employed (**Figure 2**) to understand the current (as of May 30, 2020) scenario of the countries (Afghanistan, Bangladesh, Bhutan, India, Maldives, Nepal, Pakistan, and, Sri Lanka) in terms of their confirmed and death cases ratio.

**Figure 1:**
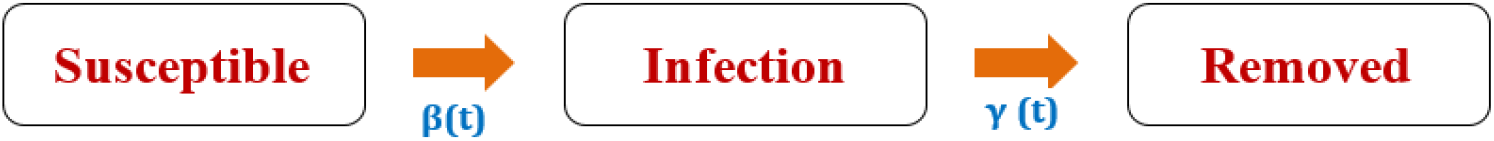
Modified SIR (M-SIR) model with the time-varying transmission

**Figure 2:**
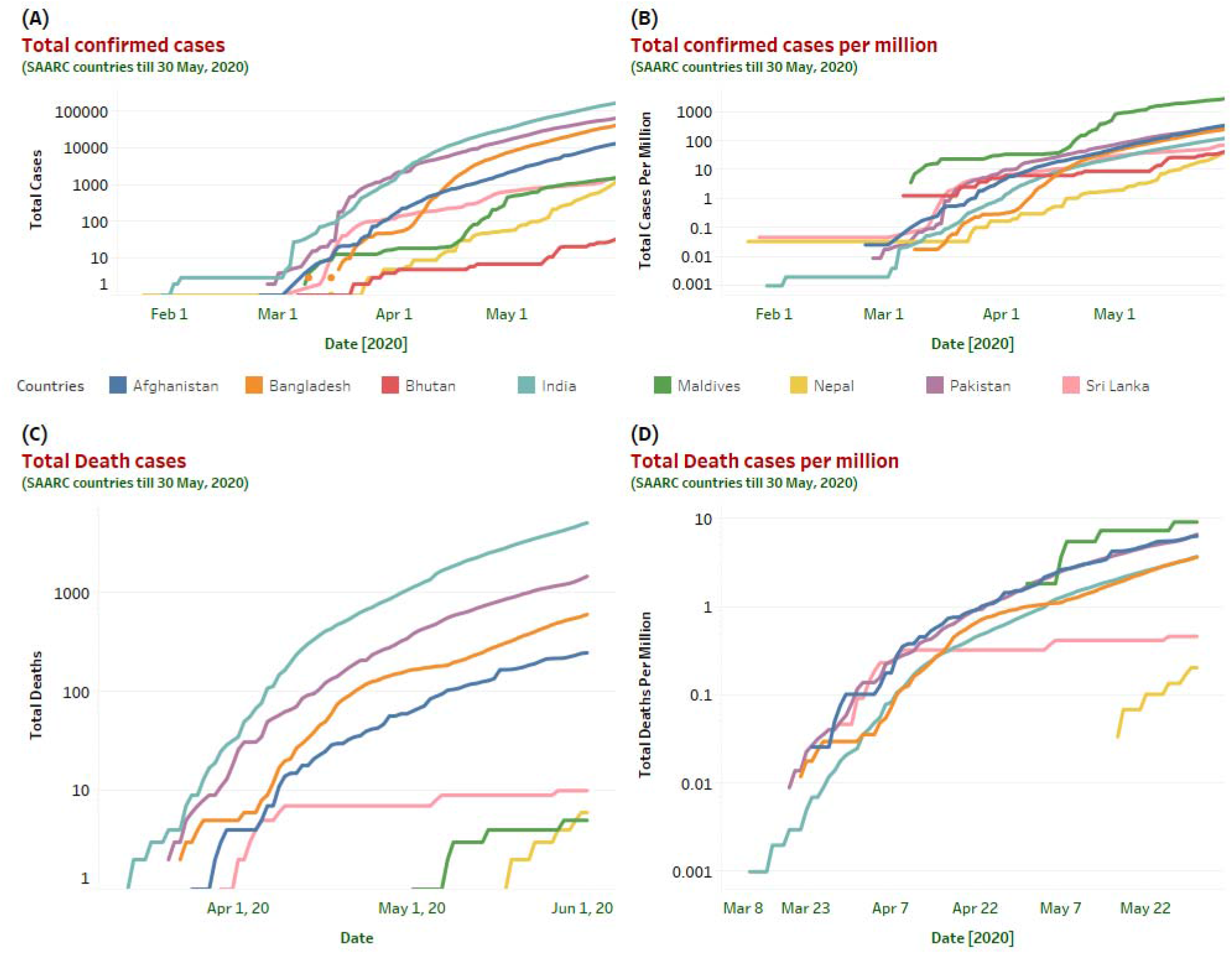
This graphical representation provides a comprehensive overview of the total number of confirmed and death cases in SAARC regions. Country-wise COVID-19 confirmed and death cases along with the number of cases per million illustrated in 2(A), 2(B), 2(C), and 2(D) till May 30, 2020. Based on the illustration on 2(A) and 2(C), India has confirmed the highest number of infected cases (173763) along with the most number of death cases (5164) in the region.

Surprisingly the slow growth of infections in the South-Asian region could be a result of a lower number of testing and testing strategy^**17**^. Initially, the testing was limited to specific individuals who have come from high-risk countries. Even their immediate contacts were also ignored primarily. The initial growth rate of COVID-19 infection in the SAARC region is comparatively lower than countries like the US, France, Germany, Spain, China, Italy, and so on. However, based on the current scenario in the SAARC region (as of May 30, 2020), India confirmed the highest number of COVID-19 reported cases. A short term model prediction for the next 20 days (till June 19, 2020) is illustrated in **Figure 3** for all the countries of SAARC regions.

**Figure 3:**
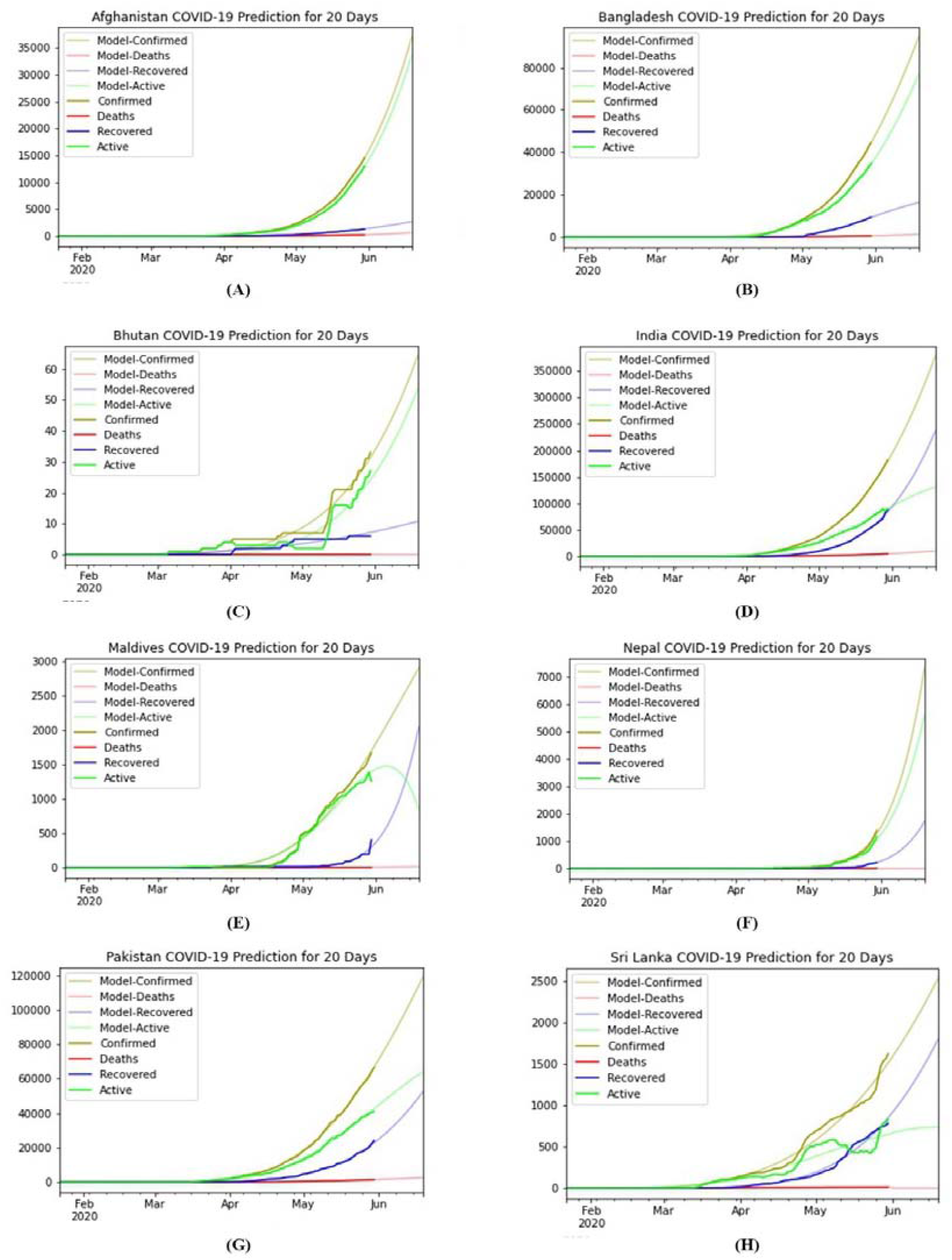
Short term prediction model for the next 20 days using M-SIR approach. This prediction provides a depiction of all SAARC countries evaluation of COVID-19 till Mid June. All the countries prediction curve are denoted here from (A-H) respectively.

Similarly, we also predicted the epidemic curve of SAARC regions for the midterm (90 days) and long term (180 days) COVID-19 cases. After predicting the 20 days of the epidemic, we noticed that India, Bangladesh, Sri Lanka, and Pakistan are increasing gradually by minimizing the active cases, and their recovery rate is also rising over time. However, in this short period, the confirmed cases of these countries are also increasing till June 19, 2020.

## 4. Discussion

The US, Italy, Spain, Germany, China are the top most affected countries in the world due to the pandemic of COVID-19. Though the novel coronavirus originated and started to spread from China but, countries from the South-Asian region are also infected faster than other countries globally. The overall population density of the countries in the SAARC regions is also too high. According to **Table 4**, we can observe the various parameters (population, land area, density, and world share) and their impact on analyzing the spread of novel coronavirus in these regions.

**Table 4.**
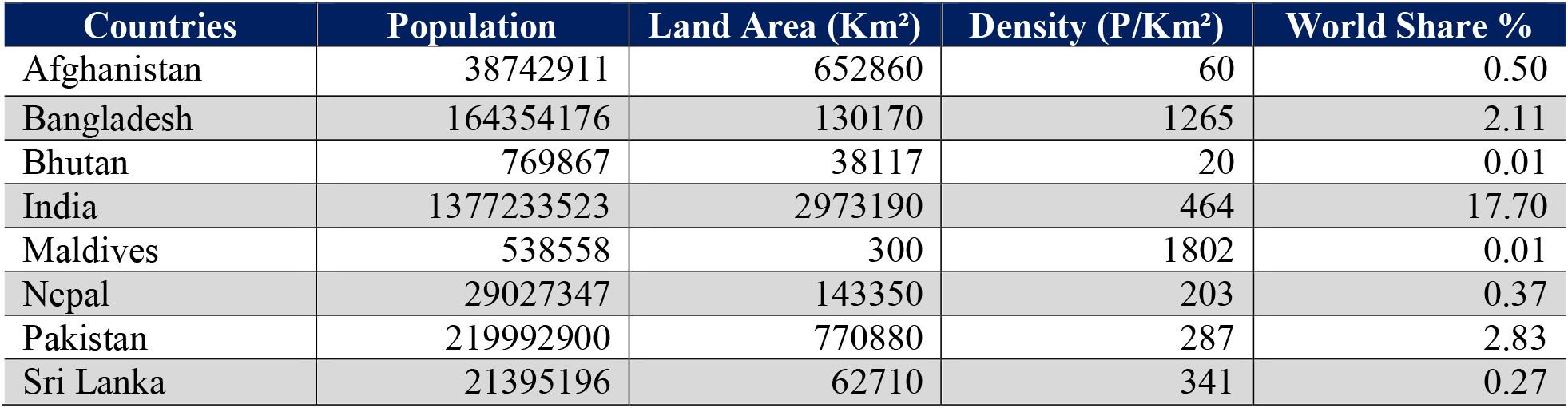
Population, total area and density of SAARC countries with their individual world share^**13**^

We further extended our prediction model for the next 90 days and 180 days, respectively. Besides, the total number of the population of a country is also parameterized because the total population of a country cannot be infected. Therefore, if we consider the total population as infected, then the probable number of infected persons remains unknown. Based on the prediction model, we tabulated the model predicted results of all SAARC countries for short term, mid term and long term prediction respectively **(Table 5)**.

**Table 5.** Predicted data based on M-SIR model for the countries of SAARC region

| Countries |  | Short term prediction (20 days) | Mid-term prediction (90 days) | Long term prediction (180 days) | Remarks |
| --- | --- | --- | --- | --- | --- |
| Afghanistan | Confirmed | 35286 | 297264 | 1092714 | Confirmed, recovery and active cases will increase at each predictive term<br>In the long-term forecast, the number of death cases will decrease comparing with the mid-term forecast. |
|  | Deaths | 652 | 5498 | 3521 |  |
|  | Recovered | 2555 | 8521 | 13416 |  |
|  | Active | 32078 | 283245 | 1075775 |  |
| Bangladesh | Confirmed | 100037 | 451840 | 867700 | significant rise will be observed in confirmed and active cases .<br>The number of recovered cases after mid-term prediction will not increase much in the long-term prediction. |
|  | Deaths | 1346 | 6080 | 11676 |  |
|  | Recovered | 17724 | 29418 | 30476 |  |
|  | Active | 80966 | 416341 | 825547 |  |
| Bhutan | Confirmed | 61 | 372 | 1683 | Large number of confirmed cases but not enough recovery.<br>The number of deaths will remain zero. |
|  | Deaths | 0 | 0 | 0 |  |
|  | Recovered | 11 | 32 | 79 |  |
|  | Active | 50 | 340 | 1603 |  |
| India | Confirmed | 378657 | 1717831 | 3709965 | The highest number of confirmed, deaths, recovered and active cases will be observed in the region.<br>It will show significant improvement in recovery cases |
|  | Deaths | 10648 | 48307 | 104329 |  |
|  | Recovered | 217681 | 1269646 | 2609589 |  |
|  | Active | 150327 | 399877 | 996047 |  |
| Maldives | Confirmed | 2873 | 5970 | 7081 | Both the confirmed and recovered cases will increase with a similar trend.<br>Death cases will also fall to zero as active cases. |
|  | Deaths | 14 | 0 | 0 |  |
|  | Recovered | 1139 | 5970 | 7081 |  |
|  | Active | 1719 | 0 | 0 |  |
| Nepal | Confirmed | 6295 | 185528 | 1089821 | Confirmed cases will notably increase. |
|  | Deaths | 0 | 0 | 2955 |  |
|  | Recovered | 1783 | 99292 | 658568 | However, the death cases will jump to 2955 from zero in the long-term prediction. |
|  | Active | 4512 | 86236 | 428297 |  |
| Pakistan | Confirmed | 118775 | 332528 | 497800 | Confirmed, deaths, recovered and active cases will increase gradually.<br>recovered cases will show almost double in the long-term prediction. |
|  | Deaths | 2534 | 7096 | 10623 |  |
|  | Recovered | 49434 | 254510 | 487176 |  |
|  | Active | 66806 | 70921 | 588234 |  |
| Sri Lanka | Confirmed | 2496 | 8832 | 23151 | In the mid-term and long-term forecast, active cases will become zero |
|  | Deaths | 0 | 40 | 106 |  |
|  | Recovered | 1840 | 8791 | 23045 |  |
|  | Active | 655 | 0 | 0 |  |

A midterm (90 Days) prediction model is further assessed until August 31, 2020, and the nature of the epidemic curve has been depicted in **Figure 4**. For all the countries of the SAARC regions, we have predicted the total number of confirmed, death, recovery, and active cases.

**Figure 4:**
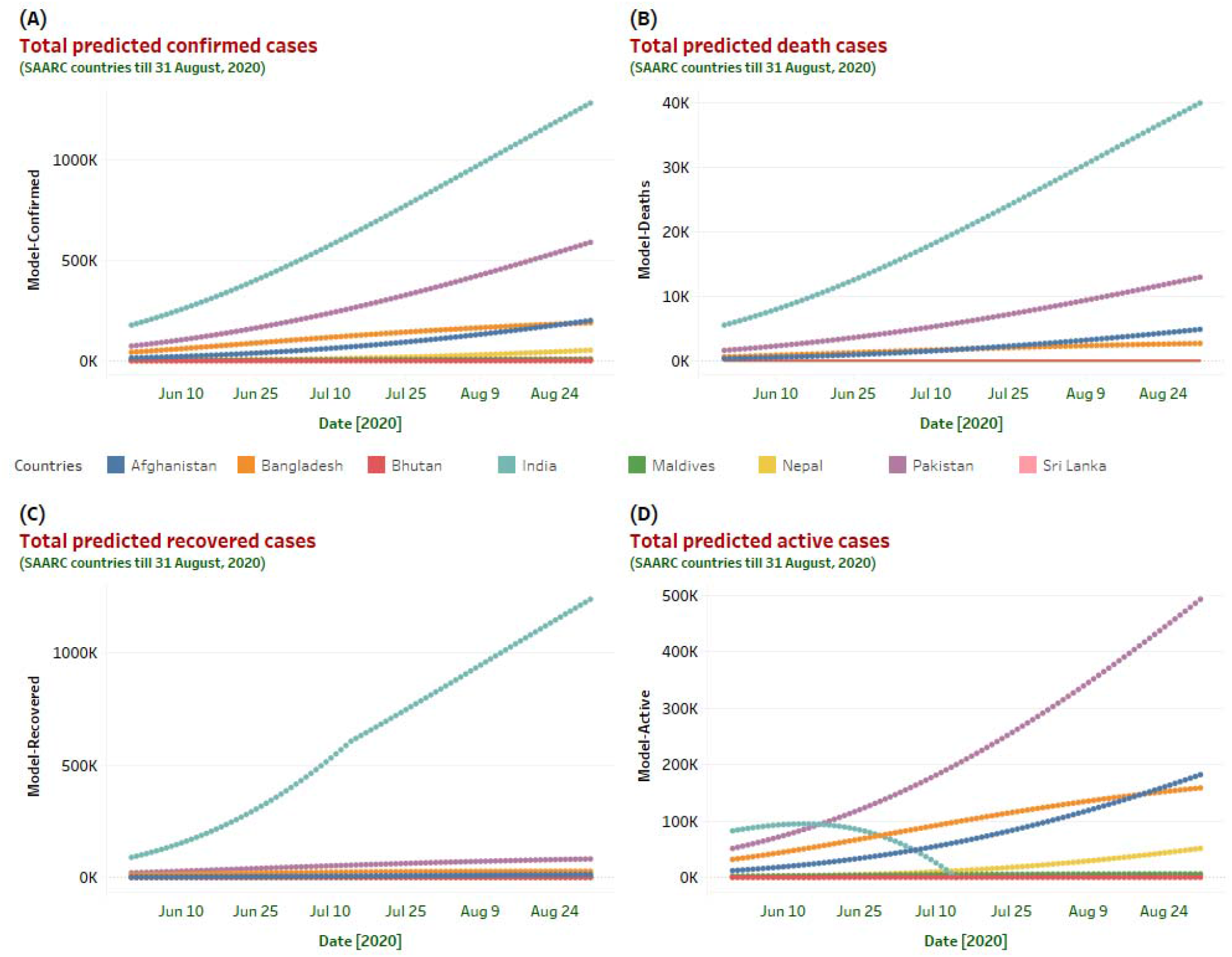
Based on the M-SIR prediction model, prediction statistics for the upcoming 90 days for all the countries in the SAARC regions has depicted here. Different predicted case scenarios (confirmed, death, recovered, and active) till August 31, 2020, helps to understand the epidemiological nature of COVID-19 in the south Asian region. The predicted model indicates that in the next 90 days prediction curve will increase sharply for India (in terms of confirmed and death cases). Moreover, India will also show a substantial increase in its recovery rate. However, the number of active cases will increase in Pakistan till August 31, 2020.

Lastly, a long term prediction model for the next 180 days (till November 30 2020) situation has been presented in **Figure 5**. According to the prediction model, active cases will fall to zero at the end of June in Sri Lanka, and at the beginning of August in India (**Figure 5**). However, Bangladesh, Maldives, and Pakistan will take more few months to reduce its active cases. On the other hand, countries like Afghanistan, Nepal and Bhutan will show a steep increase in their active cases.

**Figure 5:**
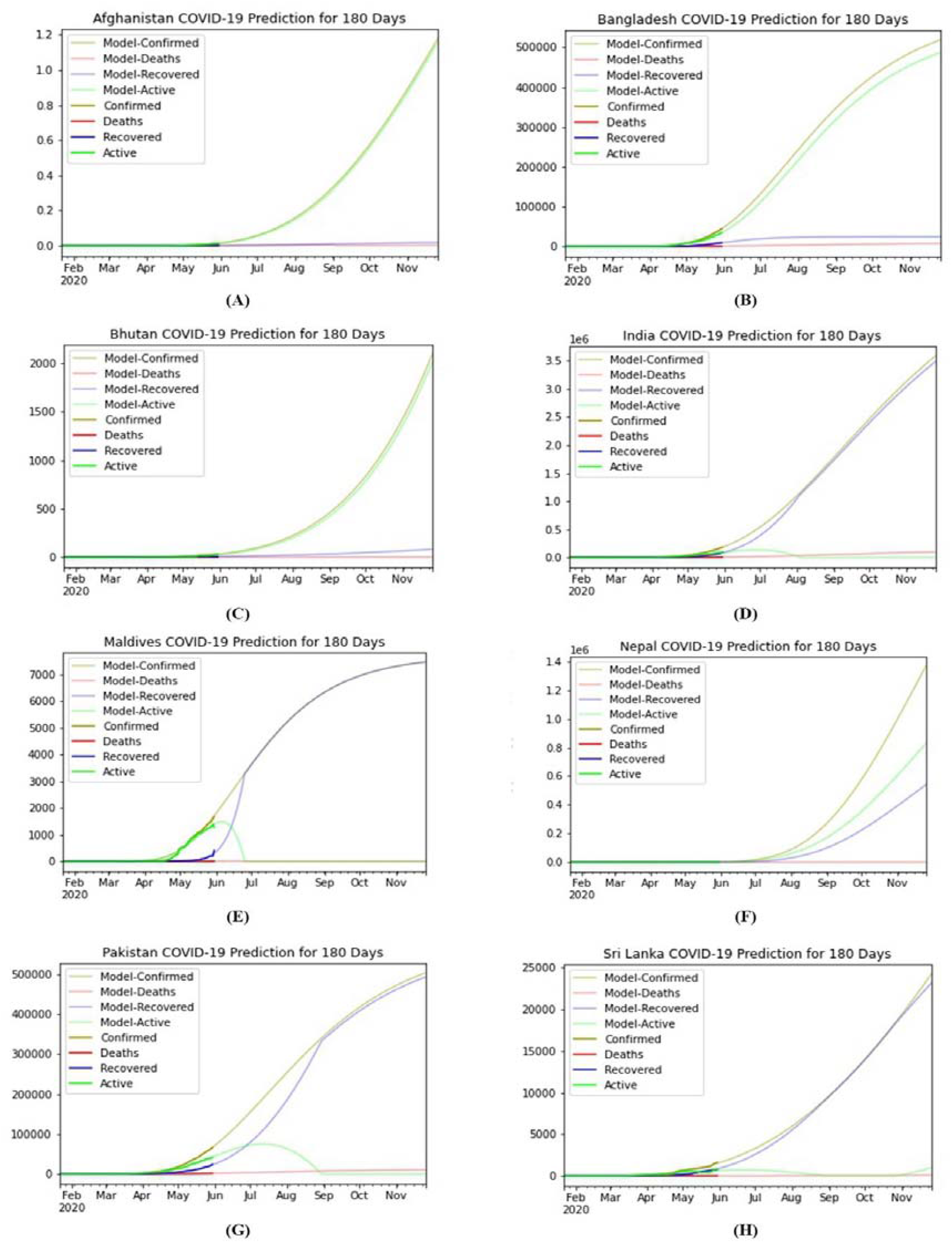
Long term prediction model for the next 180 days using M-SIR approach. This prediction provides a depiction of all SAARC countries evaluation of COVID-19 till Mid November. All the countries prediction curve are denoted here from (A-H) respectively.

## 5. Conclusion

The COVID-19 epidemic has brought unprecedented health concern for the community of all around the globe. The pandemic data of COVID-19 of SAARC countries have analyzed by using the M-SIR model to predict the confirmed, deaths, recovered and active cases with respect to the time in days. We predicted the epidemic data in the basis of short term, midterm and long term situation. To the best of our knowledge, this study is the very first COVID-19 prediction model which focused on the countries of SAARC regions. This epidemic modelling can be a helpful tool for estimating and predicting the scale and time course of COVID-19, evaluation of the effectiveness of public health interventions, and information on public health policies in SAARC countries. In future machine learning tools can be further used to identify and optimize time profile for the confinement.

## Data Availability

Not Applicable

## Author Contribution

All authors conceptualized and designed the study. SKD, KHS and MR had the idea for and designed the study and had full access to all the data in the study and take the responsibility for the predictive model and accuracy of the epidemic trend analysis with their visualization. URS and AH and contributed to the writing of the article. MR contributed to the critical revision of the report. All the visualization and data presentation methods developed by SKD, KHS and MR. All authors contributed to data acquisition, data analysis, and reviewed and approved the final version.

## Ethical Approval

Not required

